# The Association between Climate Change and Lyme Disease Incidence in Northern European Countries by the Baltic and North Seas: An Ecological Time-Series Study

**DOI:** 10.1101/2025.08.24.25334298

**Authors:** Mohammed AB Alhamadn

## Abstract

Climate change is a complex problem that often disproportionately affects global health, including its influence on microbial ecosystems, which can lead to disease outbreaks such as Lyme disease (LD). However, the extent of climate variability and its influence on the spread of the most common vector-borne disease in countries neighboring the Baltic Sea and North Sea have not been fully quantified. Therefore, this study quantified the magnitude of the (LD) burden due to climate change in the most at-risk European countries. In this ecological study, the correlation between disease incidence over the years and climate change was tested using a Spearman correlation test, and the change in incidence of LD during the period 2000-2024 was assessed using Generalized Linear Models (GLMs) with a negative binomial distribution to handle overdispersion. The study found a strong positive relationship when accounting for the tick life cycle using lagged climate variables of two years to higher rates of LD in countries bordering the Baltic and North Seas. Most importantly, a unit increase in precipitation (mm/year), after adjusting for covariates, is associated with a higher disease rate (IRR Range= 1.15-1.24). Two years of delayed temperature effect has a similar relationship (IRR range = 1.11-1.27). These findings suggest that disproportionate climate change in the region influences the spread and burden of the disease in the northern European temperate climate, mostly observed after two years of delays. Compared to previous research, this study focused on the regional impact of climate change on microbial life and disease spillover, which causes public health problems. There is an urgent need for a collaborative and comprehensive program that includes environmental, human, animal and vector data factors for future research to aid in disease control.

## INTRODUCTION

Climate change, the continued alteration in the average norms of temperature and other weather-related variables, has been a core and complex burden on global health over the last few decades. Recent evidence indicates that there is an increase in warming of 1.25 °C every 10 years ^1–5,14^. However, these changes are not homogeneous across the globe; they are proportional, and each region has its own rate of change. New evidence indicates that Arctic warming has shown even faster odds of warming, up to four times, compared to warming in other places around the world ^23^.

Researchers have examined the connection between rapid Arctic warming, that is, Arctic amplification, and the Baltic Sea region and found a strong atmospheric connection between them ^31^. According to the European Environmental Agency, the Arctic and Baltic Sea are directly impacted by climate change, ice cover has hit its lowest points in recent years, and it has increased the annual precipitation and water temperature. Both Arctic and Baltic sea ice are observed to be melting rapidly, which has not been seen before. Climate modeling has projected an imminent ice-free event by 2050, which is attributed to the impact of climate change on the area, making the European region the most influenced by global warming ^30^. Linear climate trends between 1982 and 2012 showed a strong correlation between changes in sea surface temperature in the North and Baltic Seas, emphasizing their interconnected behavior ^39^. To further explore this, data from to 1980-2022 in Europe found that warming has uneven impact rates in all its regions ^18^. Their study argued that the northern part of the EU is impacted the most by shifting climatic conditions, meaning that the shift from colder to warmer conditions is faster than that in other areas of the EU, linking it to Arctic amplification ^20,23,29^. In addition, another study confirmed the findings in northeastern countries in Europe and in a wider scope, such as France, which experienced an extreme weather event in 2019 at 46.0 °C, confirming the wider impact of this occurrence ^17^.

This rapid disproportional change in climate factors per decade acts as a stressor and disturbs the equilibrium and interrelationship between environmental, human, and animal health ^5–11^. Direct divergence in the influence of what was once normal to ecosystems leads to new ways for microbes to spread and to find a new home or pathway for disease transmission ^13,14,30^. A wide range of pathogens are influenced by this phenomenon, one of which is Lyme borreliosis ^32^.

Lyme disease (LD) is a zoonotic vector-borne disease that is transmitted from ticks to humans through a tick bite ^1^. LD was first noticed in 1909 in Europe and was then discovered in Lyme, Connecticut, in 1983. This discovery began with an outbreak that occurred in 12 communities and infected 43 people in 1975^5^. Later, the bacterial causative agent (Borrelia burgdorferi) was isolated from a tick, formerly known as Ixodes dammini, now called the blacklegged tick ^2^. There has been research on alternative methods of transmission; however, these claims have not been scientifically supported. Currently, LD does not have an approved vaccine for humans ^6^.

Several outbreaks have occurred in Europe and the United States over the years, causing an increase in global health threats. LD is considered to be the leading burden of all vector-borne diseases ^3^. It causes a wide range of symptoms that start as mild symptoms, such as fever, rash, and pain, and then progress to severe symptoms, such as inflammation of the joints, neurological, and heart conditions ^4^. It is estimated that the annual prevalence of the disease in Europe is more than 200,000 cases each year, and about 476,000 reports in the USA per year ^3,7^. The most common ticks that carry the disease are Ixodes scapularis and I. pacificus in the USA, and I. ricinus and I.persulcatus in the EU ^3^. Animals have also been reported to become infected ^8^.

Ixodes ricinus, the most common tick in the EU, spends most of its life cycle outside of a host, and it takes approximately two years to complete its full biological form, where it can survive for up to six years depending on weather conditions ^3,8,11,12,15^. They rely on the active presence of water to survive, and their movement is completely influenced by temperature ^10,26,27^. Moreover, ticks thrive in mixed forest settings with high humidity levels. Tick behavior (I. ricinus) in seeking hosts is associated with humidity and higher temperatures. Studies have reported that the northern (and western) areas of the EU have favorable living conditions for ticks, focusing on areas where the temperature is at a constant 6°C and the relative humidity is higher (above 80%), which is a result of higher annual precipitation ^13–16^.

Adult ticks acquire the causative agent from infected animals during the nymph stage of their life cycle. When a tick is infected, the bacteria live in the tick’s gut in a dormant state until activation, which occurs approximately 24 h after biting and attachment to the host ^11^. Studies have shown that once an infected tick feeds on a blood meal, the bacterium travels to its salivary glands and remains inactive until it can pass from the tick to the host and cause infection ^9–13^. Previous evidence suggests that while tick’ behavior is influenced when carrying the bacterium ^38^, some different pathogens have a mechanism that improves their ability to infect hosts and makes transmission even more efficient ^12^. An example of this is the production of Outer Surface Protein C (OspC), a bacterial protein that helps Borrelia spread from ticks to vertebrate hosts during feeding ^12^. An experimental study showed that this process could take less than 17 h because of the protein expressed by the bacterium for it to move from the tick to the host ^9^; however, it was not scientifically backed in human hosts.

The role of animals serves as an additional layer of this complex problem. Ticks utilize several animals to move and control the areas where they are endemic ^10–15^. Studies have found that ticks have a selective preference for the animal hosts they use as reservoirs. Ticks tend to feed on certain large mammals (e.g., deer) around the world. They are less likely to acquire infections from these hosts; however, deer are associated with tick movements. In the EU, rodents (e.g., wood mouse and bank voles) serve as important hosts for smaller species. In addition, it has been suggested that the disease can quickly spread to a new area using birds and that localized transmission occurs through mice or deer ^8–11,15,29^.

Factors related to tick ecology and disease events have been investigated for many years. Studies have found that there are cues that facilitate disease spillover and its spread through zoonotic cycles. These factors are related to tick populations, host availability, location, and seasonality and climate variations ^1,26,29^. However, research has overlooked the interconnection of all these factors as contributing to a higher risk of LD in northern EU regions. The presence of all these factors, especially for vulnerable countries in the north of the EU ^17,18^, makes the layer of complexities seen at a higher rate than their interaction elsewhere. It is important to model climate and disease incidence while accounting for tick biological terms; therefore, the corresponding author created lagged variables for delayed climate influence to model delayed climate influence based on the tick life cycle that can take two years to become in a questing behaviour ^9,11^, leading to an increased risk of disease across countries over time.

This study aimed to unpack this complex problem through a model that quantifies the relationship through measurable variables related to the disease, time trends, and environment, and its delayed effect on the increased rates of LD in the coastal regions of Europe. Several variables were collected to test 1) whether the average change in climate variables over the years correlates with increased LD incidence rates, 2) determine if the delayed lagged variables (one and two years) to temperature and rainfall are associated with a higher disease risk, and 3) assess the disease dynamics affecting LD incidence in the northern region of the EU from 2000 to 2024. At this early stage, the hypothesis is operationalized by evaluating how interconnected variables relate to climate in northern countries, and how these associations influence disease incidence when biological factors are incorporated. An ecological study with a time-series design covering population-level data over time in northern regions is needed. The outcome would provide a scientific basis for this hypothesis and guide additional research to better correlate climate variables with disease burden and apply appropriate intervention programs to mitigate higher rates of disease.

## MATERIALS AND METHODS

### Study Design and data collection

This ecological study aimed to test the association between climate-related factors and Lyme disease incidence from 2000 to 2024 in several Northern European countries. The study included yearly data to understand the regional association at the macro level for the variables; therefore, all data used were aggregated. Nine European countries were included in this analysis, and these countries were considered based on 1) their utmost climate exposure, that is, their geographical location in the Baltic Sea or the North Sea, and 2) data on disease case count or rate availability for at least four years to perform lagged effects analysis.

Regarding the climate exposure parameters, all data extraction used an Application Programming Interface (API) based on 1) latitude and longitude, and 2) time in years. Meteorological exposure variables were obtained from the National Aeronautics and Space Administration (NASA) Langley Research Center (LaRC) Prediction of Worldwide Energy Resources (POWER) during the analysis to obtain the yearly average temperature (T2M) and precipitation (PRECTOTCORR) data. NASA’s POWER Project (retrieved May 10, 2025), dataset for POWER Daily 2.x.x, at https://power.larc.nasa.gov. The yearly average temperature (°C) and total precipitation (mm/year) were calculated by averaging daily values over the course of a calendar year, with precipitation totals adjusted for 365.25 days. Furthermore, to account for the biological plausibility of ticks becoming infectious, two lagged covariates were created for each climate index (T2M_lag_1_year and T2M_lag_2_year) for average temperature and (Precep_lag_1_year and Precep_lag_2_year) for average precipitation, totaling four additional mean climate elements. Similarly, the population size for each country was collected from the World Bank Statistics using the same extraction method and (wbstats) package. All these data were then merged into one large dataset during the analysis phase.

The outcome variable is the annual incidence of Lyme disease, which was obtained from the Hopkins Lyme Tracker https://www. hopkinslymetracker.org/explorer/. Data availability for the disease is over the period of 2000-2024, no restriction of date has been placed in this study, but three countries (the Netherlands, Denmark, and Sweden) have been dropped from the analysis due to missing disease data. Disease rates are presented as rates per 100,000 people in this study.

In the descriptive statistics table, Figure 1 shows the nine countries included in this study. The outcome variable is presented as the overall raw total number of cases per country. Climate metrics are presented as the average per year for temperature in °C (SD), and precipitation is averaged in mm/year (SD). The country’s mean population shown is an average census for the population in which the country participates.

**Figure 1:**
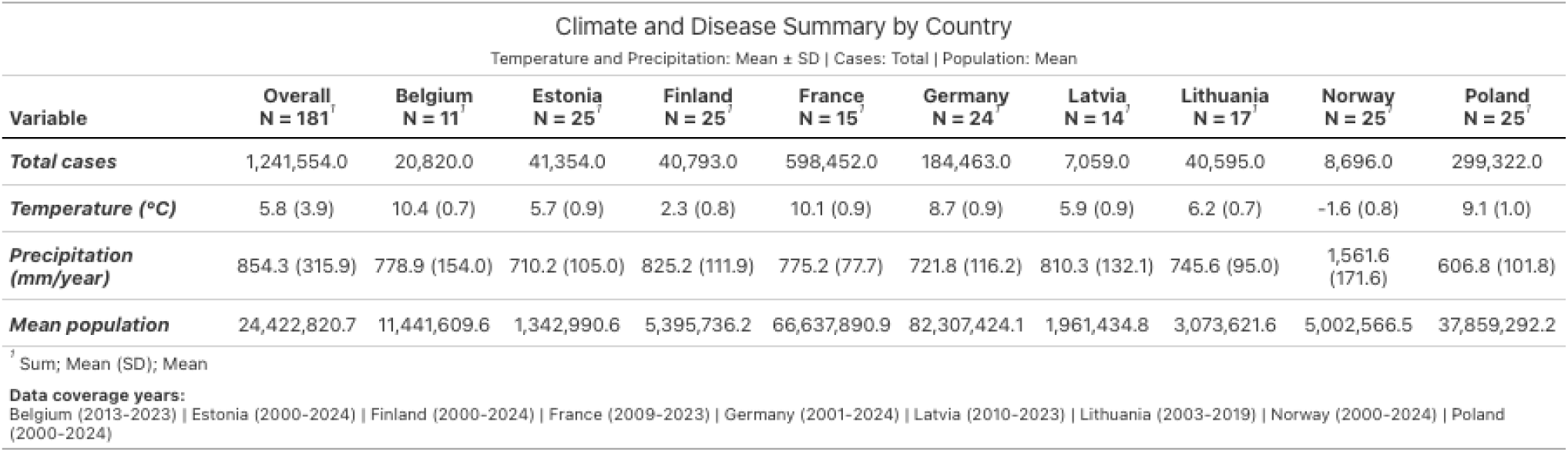
Descriptive summary table.

**Table 1:** Model definitions, variables, and fit across countries.

| Model Name | Definition | Variables | Number of parameters (K) | Best fit for (n) of countries |
| --- | --- | --- | --- | --- |
| Model1 | Core environmental measures + population control | Temperature, precipitation, log(population) | 4 | – |
| Model2 | Core + population control + one year lagged effect | Temperature, precipitation, temperature_lag_one, precipitation_lag_one, log(population) | 6 | – |
| Model3 | Core + population control + one year lagged effect + covariate control | Temperature, precipitation, temperature_lag_one, precipitation_lag_one, log(population), year | 7 | – |
| **Model4** | Core + population control + two year lagged effect + covariate control | Temperature, precipitation, temperature_lag_two, precipitation_lag_two, log(population), year | 7 | 9 out of 9* |

\* Model 4 best fit in all countries

### Statistical analysis

This study involved a complex relationship between climate variables, disease incidence rates, and time. Here, a Spearman correlation test was used to assess the correlation strength of the variables of interest. Due to the biological aspect of ticks, two lag-year variables were added to assess the delayed correlation for one and two years to the frequency of LD incidence over time. Subsequently, General Linear Models (GLMs) were used to assess the association between climate metrics and increased case incidence, while accounting for covariates such as lagged variables, population, and year as offsets. After using Poisson Regression, the assumption was not met and was alternatively shifted to using Negative Binomial Models fit, to account for the observed overdispersion. Model selection was assessed using the Akaike Information Criterion (AIC) for model comparison, the Variance Inflation Factor (VIF) for multicollinearity, and Pearson residuals for the goodness-of-fit test. These tests revealed that Model 4 is the best model of fit with VIF results below 3, (AIC) values ranging from 116 to 399, which are the smallest AIC values in all nine countries, and (1.8 dispersion) residual values centered around zero. The model variables and selection align with the biological traits of the vector, which can become a threat. In the final model, ‘population’ was included as an offset to account for demographic change over time, a better comparison unit to changing rates over the years and within the countries. The variable (year) was added as a covariate to control for time-trend variations in reporting, intervention, and disease ecology changes. Therefore, the sequential models tested are as follows:

## RESULTS

Figure 2 shows that the rate of Lyme disease has increased over the years in the EU, with the disease rate increasing by up to 3-folds. The rates were the highest and consistently increased in countries near the Baltic Sea (Estonia and Lithuania) during the study period (2000-2024). The number of new LD cases has also increased sharply in countries that are coastal to the northern and western regions of Europe, such as Poland, Germany, Belgium, and France. The findings are consistent with two time trend studies by *Skufca* ^33^ in Germany and *Paradowska-Stankiewicz* ^34^ in Poland.

**Figure 2:**
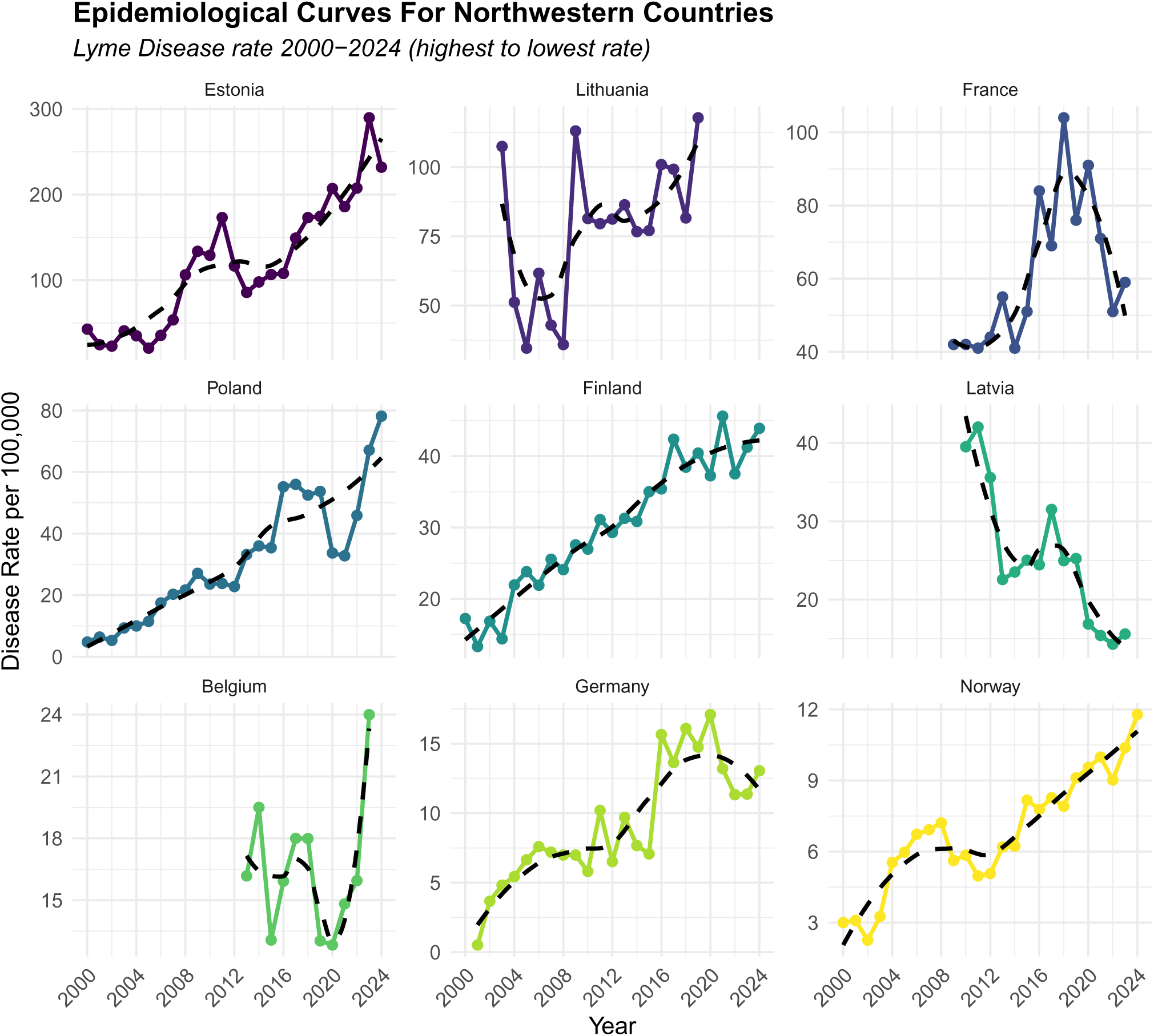
Epidemiological curve showing LD rates over the years (2000–2024).

The univariable analysis in Figure 3 (Negative Binomial Regression) was consistent with the results of the Spearman Correlation Test when testing each relationship alone on the outcome. A 4-10% positive amplified relationship is observed in seven of the nine countries for year trends. The tests also revealed that Lyme disease (LD) incidence rates and environmental variables (current and lagged years) were positively associated in all countries. Unadjusted IRRs showed that a one-unit increase in the current year’s average temperature in Estonia, Poland, and Germany was associated with an increase of 26-44% in LD incidence. An average unit rise in temperature lagged by one year was associated with 15-42% of cases of LD in Finland, Estonia, Poland, Germany, and France. Additionally, one unit increase in the average temperature two years before, is estimated to be associated with a 21-36% increased in cases in Estonia, Lithuania, Germany, and France.

**Figure 3:**
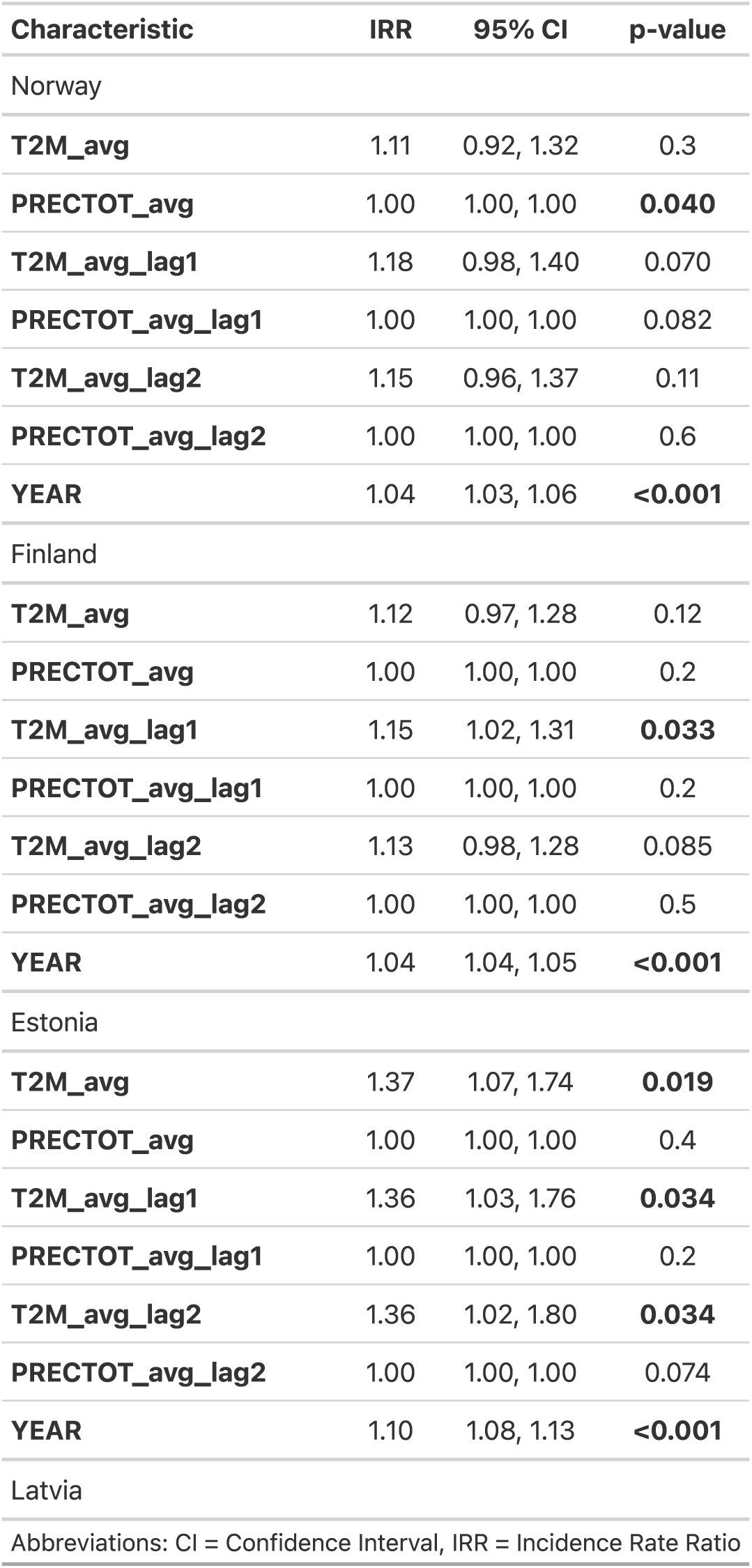
Univariable Analysis (First Page Shown). The complete table is available in the Appendix.

After adjusting for covariates in Model 4, which was selected based on the best model fit, most countries showed an independent increase of 3-12% in cases annually (Figure 4). Central to this discussion, the results indicate that lagged environmental variables of two years have a significant association with LD in six out of the nine countries. For example, an average increase in one unit of precipitation that occurred two years prior was strongly associated with a higher disease incidence in Estonia (IRR= 1.24, 95% CI: 1.05-1.48), Lithuania (IRR = 1.15, 95% CI: 1.03-1.28), Belgium (IRR = 1.21, 95% CI: 1.17-1.26), and France (IRR= 1.22, 95% CI: 1.06-1.41), p=<0.05. In addition, a strong association was observed in three countries for the same duration of delay from the average temperature to disease incidence. Data showed that Norway (IRR=1.11, 95% CI: 1.02-1.21), Germany (IRR=1.13, 95% CI: 1.01-1.25), and France (IRR= 1.27, 95% CI: 1.05-1.53), p=<0.05, observed an increase in disease incidence when there was a change in average temperature. Furthermore, the change in average precipitation in the current year by one unit was significantly associated with the outcome only in Estonia (IRR= 1.25, 95% CI: 1.10-1.43, p=<0.05). However, no country showed an association between the change in the average temperature of the same year and the risk of disease after the final selection of the model and the control of covariates.

**Figure 4:**
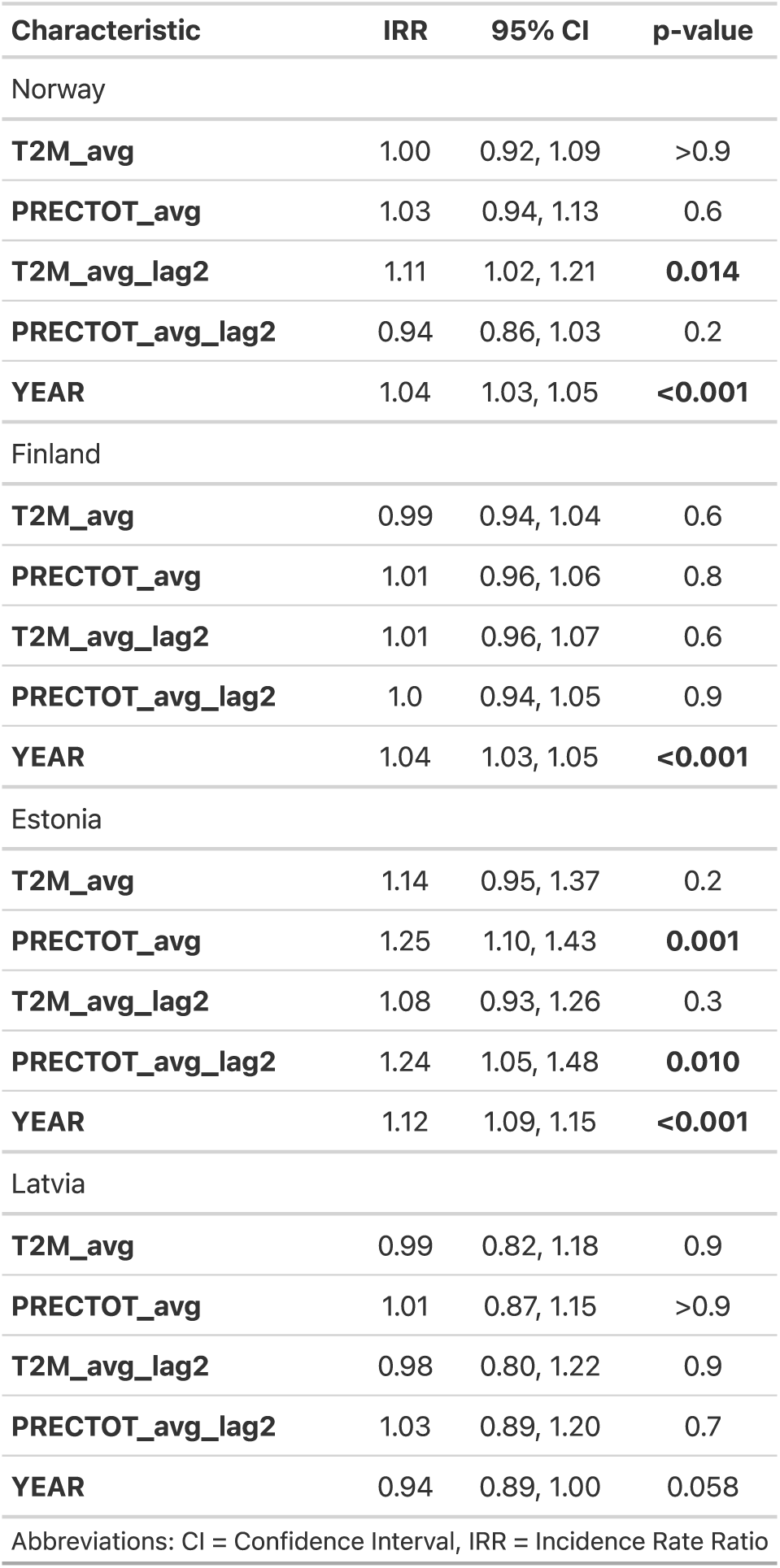
Multivariable Analysis Using Negative Binomial General Linear Model (First Page Shown). The complete table is available in the Appendix.

These findings indicate an independent association between climate change variables and Lyme disease incidence in Northern Europe. That is, when there is an average increase in environmental factors by one unit in countries bordering the two seas, it is expected to experience a higher rate of cases two years later, which is attributed to the influence of climate. The delayed precipitation effect was the most significant variable for the increase in the number of LD cases. Hence, Estonia has been the country most burdened by Lyme disease (LD) over the years due to climate change. These findings further support the initial hypothesis that climate change contributes to a higher number of LD events by affecting ecosystems unequally, leading to a higher risk of disease in several northwestern European countries, a region with a significant climate impact.

In addition, the overall LD rate in this region has been progressively increasing since 2000. The spatial distribution of Lyme disease has shifted from one country to another, which is highly associated with climatic variables. In figure 5, the map suggests that the incidence rate has shifted geographically from Scandinavian countries to the Baltic states and then to other neighboring countries, where a higher number of cases were found. These findings align with the author’s hypothesis that worsening climate conditions over time correlate with changing disease dynamics and increasing spillovers in the northern EU region.

**Figure 5:**
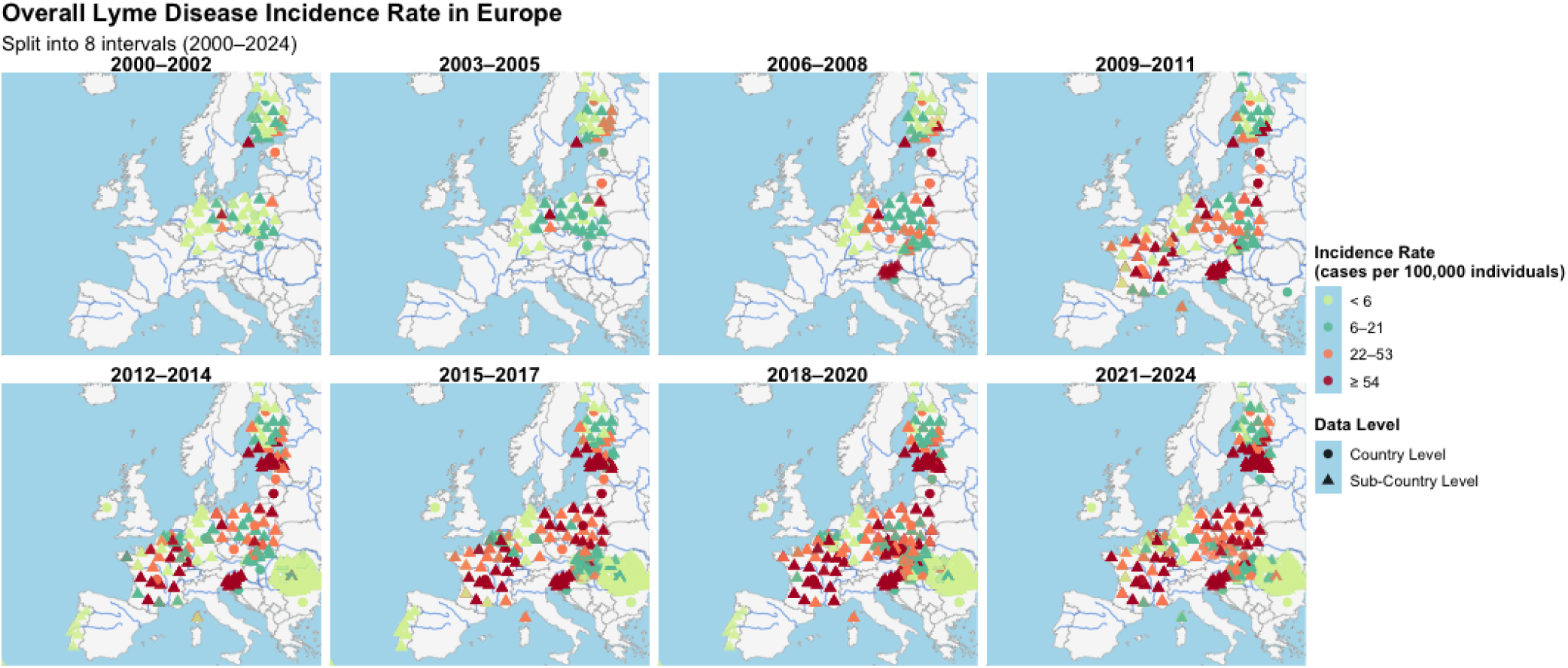
Overall Lyme Disease Rates from 2000 to 2024 in Europe. Data from Johns Hopkins Lyme Tracker (hopkinslymetracker.org).

## DISCUSSION

This multicountry ecological study was conducted to explore the association between climate change in places near the Baltic Sea and North Sea in Northern Europe and Lyme disease incidence from 2000 to 2024. Europe is known to experience microclimatic distortions, mostly in the north, and tremendous biodiversity differences, with a higher burden of Lyme disease over the years. The methodology of this study was used to quantify the risk of disease from the influence of climate in that region of Europe during the study period. To date, research that integrates environmental units into human case risks is lacking. While the majority of LD studies have performed temporal year trends, in this analysis, more granular connections were considered in relation to the data on environmental and population changes from 2000 to 2024. This analysis aids in the understanding of the higher incidence of LD in the EU. It is necessary to explore remote sensing data variables related to climate to study the increase in disease rate in the northern region of the EU.

The hypothesis in this paper is to explore whether the coastal northern countries in the EU, which face changing climate factors the most, are unequally influenced by the burden of disease. This research highlighted that there is an association between climate change and increased LD risk in the included countries in Europe. The burden of the disease is uneven and depends on the region and variations in the climatic conditions. The disease has spread rapidly over the years, with higher rates attributed to the changing climate in countries bordering the Baltic Sea and its linked maritime climate. This study found that as Arctic amplification occurs, so does microbial amplification, and this pronounced ecosystem change is now being observed from the northern borders of the coastal regions.

These results are consistent with the biological plausibility of Lyme disease vectors. Model four, which included two-year lagged variables for temperature and precipitation, was the best-fitting model in this analysis. This supports the idea that transmission dynamics are attributed to environmental changes when accounting for tick life cycles. As a result, changes in temperature or precipitation in an average year in the EU have an impact on disease incidence patterns two years later. Thus, the disease rate changes over time in countries considered to be at risk. Figure 6 shows the average estimated disease rates for the two time interval 2000-2011, and 2012-2024. There is a clear increase in estimated rates in the northeastern portion of Europe, where higher rates are observed in countries closest to the Baltic Sea, making the effect of climate change on the Arctic to regional waters, such as the Baltic Sea, and the changing zoonotic disease ecosystems as a result of this region being a strong argument. While accounting for the biological plausibility of the vector, the disease expansion is growing at a faster rate. Consequently, while investigating these remarks, the countries with the highest disease incidence rates for the longest period were Estonia, Lithuania, Poland, Finland, Germany, Norway, and France (Figure 2). This highlights the need for improved monitoring and intervention strategies to address the regional epidemic burden.

**Figure 6:**
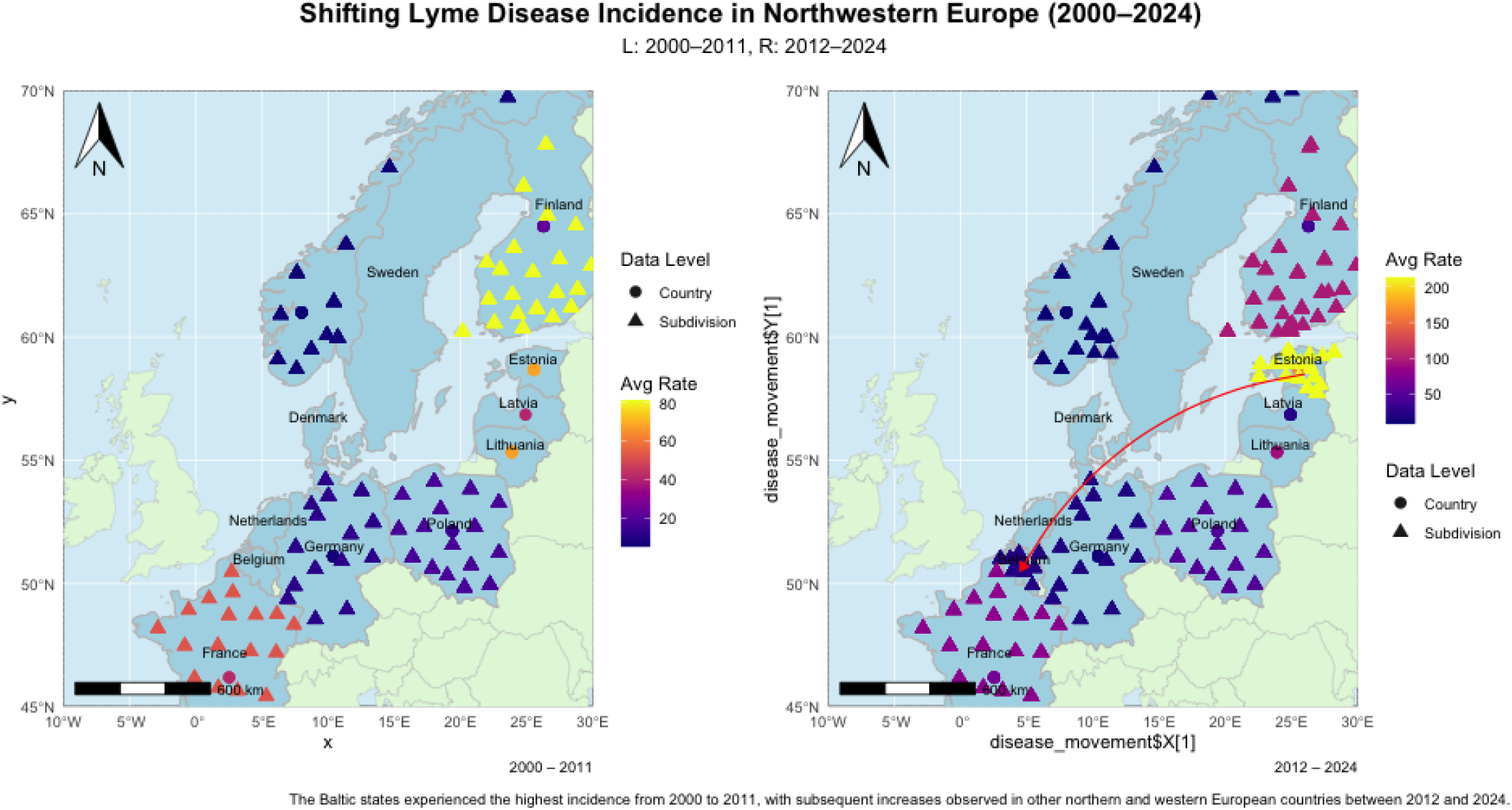
Mapping Disease Movement: Lyme Disease rates in Northwestern countries (2000–2024)

The United States has reported similar findings with respect to the link between climate and Lyme disease incidence ^29^. For comparison, data rates from the USA were also analyzed through the same pipeline at the end of the study to contrast the risk rate differences of the disease. After controlling for covariates, the incidence rates and disease burden were significantly higher in Estonia (IRR = 20.3), Lithuania (IRR = 13.4), and several other countries than the disease rate in the United States during the study period (Figure 7).

**Figure 7:**
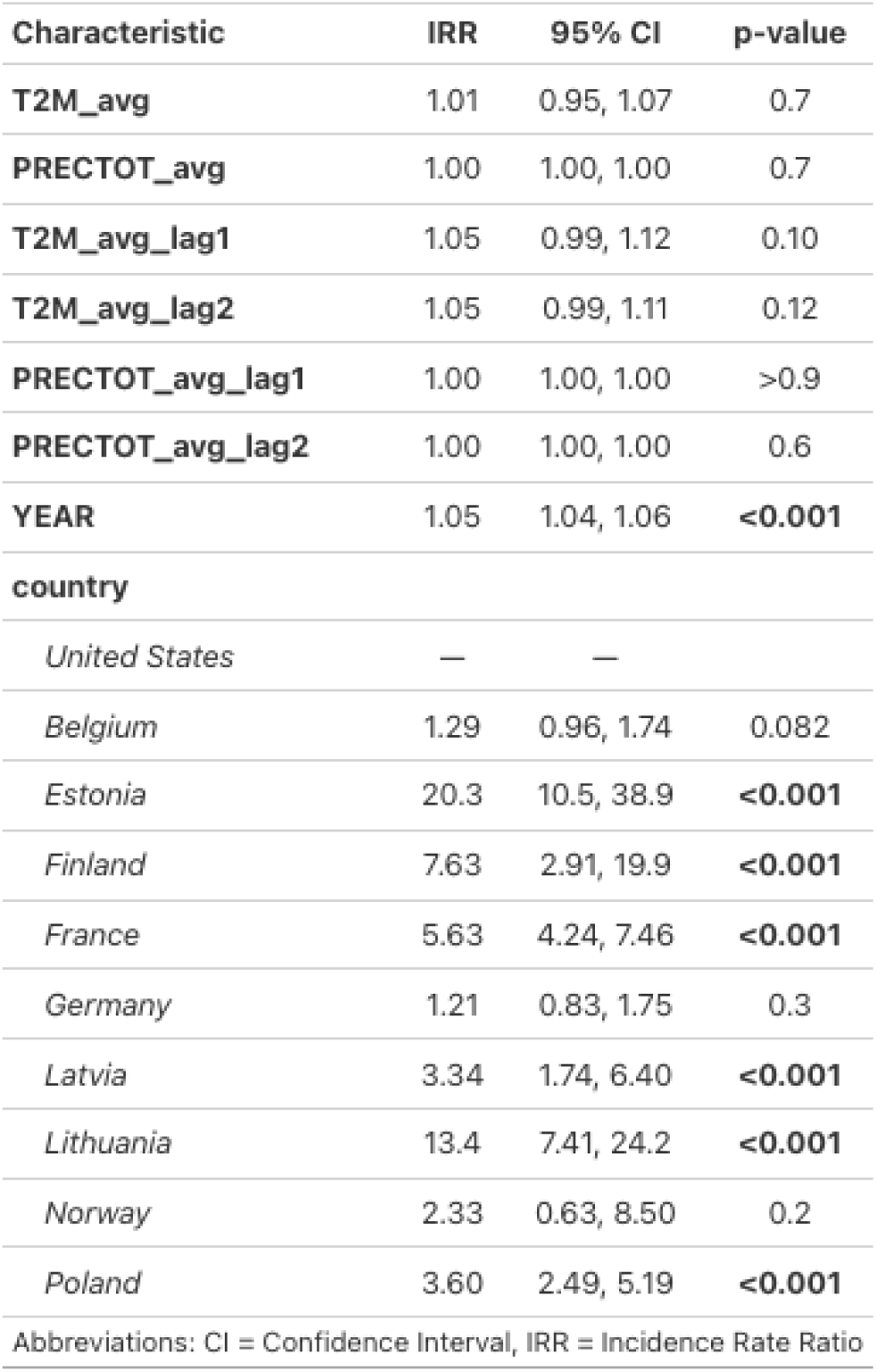
Estonia and Lithuania had about 20 times and 13 times the incidence of United States, respectively.

Lyme disease is endemic to other European countries in the central and southern regions of Europe. The scope of this study is to investigate the frontline states of the changing climate using a method that can quantify arctic amplification through disease incidence in the northwestern part of the EU. This study discussed the hypothesis and found a clear strong association between climate and disease incidence in countries near the two seas at a faster speed, suggesting with concurrent research that an environmental change to case surge could take up to 2 years, which might explain the fluctuation and delay of events based on regions. Estonia and Lithuania showed the strongest relationship between climate change variables and increased incidence over time. Disease rates are estimated to be up to 20 times higher than those in the United States. Hence, with disproportionate climate change, there are short and long effects, those effects are associated with how fast or slow the disease spreads.

### Strength and Limitation

The strength of this study is the use of advanced statistical modeling to explore the association between climate and Lyme disease incidence in a time-series ecological study design. Statistical analysis assessed several models before fitting them to the data. A negative binomial general linear model was used to account for data overdispersion. The results of the final model showed a positive association, accounting for covariates. Moreover, more meaningful results were obtained when exploring lagged variables (temperature and precipitation) with the incidence rate of the disease. Several countries were analyzed while accounting for the country’s heterogeneity, elucidating the hypothesis that ongoing trends affect more areas in the region.

There are several possible biases due to the nature of the study design and data collection methods. Ecological studies are often prone to several epidemiological concerns. First, the study exposure and outcomes were measured at the population level from 2000 to 2024, not at the individual level. Thus, there is a possibility of ecological fallacy in this study; the interpretations for each country might not be applied to small regional or personal impacts. Individual-level data are needed to confirm these findings and address the existing research gaps. Second, to account for temporal bias, the study included variables that account for the biological plausibility of the climate on the tick (one and two-year lagged). Additionally, to address differences in temporal trends between countries, a multivariable negative binomial regression model analysis was used while adjusting for time and population size.

Selection bias is a potential threat to the validity of this study. A threat may arise from the countries excluded from the region or the number of years of data available for them. First, the analysis excluded three major countries that were originally intended to be included: the Netherlands, Denmark, and Sweden. The original dataset lacked case incidence data for these countries. Additionally, not all countries contributed equally to the study (2000-2024). Some countries contributed more than others did. Collectively, these two issues could introduce a selection bias into the study.

Uncontrolled confounders can also bias the analysis if they are not adjusted for. The aim of the analysis was to assess the relationship using a parsimonious model while avoiding overfitting. The country’s healthcare and socio-economic status, and the definition to diagnose and report LD confirmed cases are not alike and vary over time, both between and within countries. To address this, the best viable approach was to use data from a reputable source, the Johns Hopkins database, in which the data were extracted from each country’s public health agency, and then using the variable (year) to account for year-to-year changes within countries. Nevertheless, this does not completely remove the reporting and diagnostic bias that could arise from individuals seeking care; thus, underestimating the true case numbers is still possible. Regrettably, there are no data to directly test animal factors in the proposed models. And, geocoded data on tick populations, as well as their infection status, were not available during the analysis. The absence of these variables indicates possible confounding factors in the study.

Research has linked climate change to an increase in the number of infectious diseases with zoonotic ability globally, including Lyme disease ^35^. However, this study examined the correlation between changes in climate units in climate-sensitive regions and the high prevalence of cases in the Upper EU region. The dynamics of zoonotic disease transmission are shaped by changing ecosystems through the influence of climate change. To better understand this relationship, it was important to assess (LD) risks in an approach that considers how climate influences disease events. Finally, it remains unclear how microbes interact within their ecosystems and how animal factors contribute to higher tick densities and movement. The role of vegetation change and land use, genetics, and other variables in persistent Lyme disease counts are all possible factors that have not been investigated here ^10,12^. More research is needed to understand how tick density and infection rates are affected by intrinsic factors such as genetics and microbiome competence, extrinsic factors such as host availability and emerging hosts ^20^, and other climate variables that influence tick density and infectiousness.

### Recommendation

Europe lacks a coordinated and standardized surveillance system, which has previously caused issues such as the misidentification of tick species ^37,38^. Therefore, strong integrative collaboration is needed to improve sampling and reporting systems. This effort can be carried out using a One Health approach. Reporting data across all domains requires interdisciplinary teams, including those that deal with environmental and vector conditions (biologists), animal roles (veterinarians and animal health practitioners), and standard confirmed human infections (healthcare professionals). This is necessary not only for disease tracking, but also for improving analytical accuracy and generalizability to understand how climate change affects microbial ecology.

## Data Availability

All data supporting this study are available from public sources:
- Human disease data: Johns Hopkins Lyme Tracker (https://www.hopkinslymetracker.org)
- Climate data: NASA POWER Project (https://power.larc.nasa.gov)
- Population data: World Bank Statistics (https://data.worldbank.org)

https://power.larc.nasa.gov

https://data.worldbank.org

## AUTHOR CONTRIBUTIONS

The corresponding author oversaw the entire study, from conceptualization to methodology, data analysis and writing.

## AUTHOR COMPETING INTERESTS

The author has declared that he has no conflicts of interest.

## Appendix

### Supplementary Tables

Complete tables referenced in the main text.

**Table A1:**
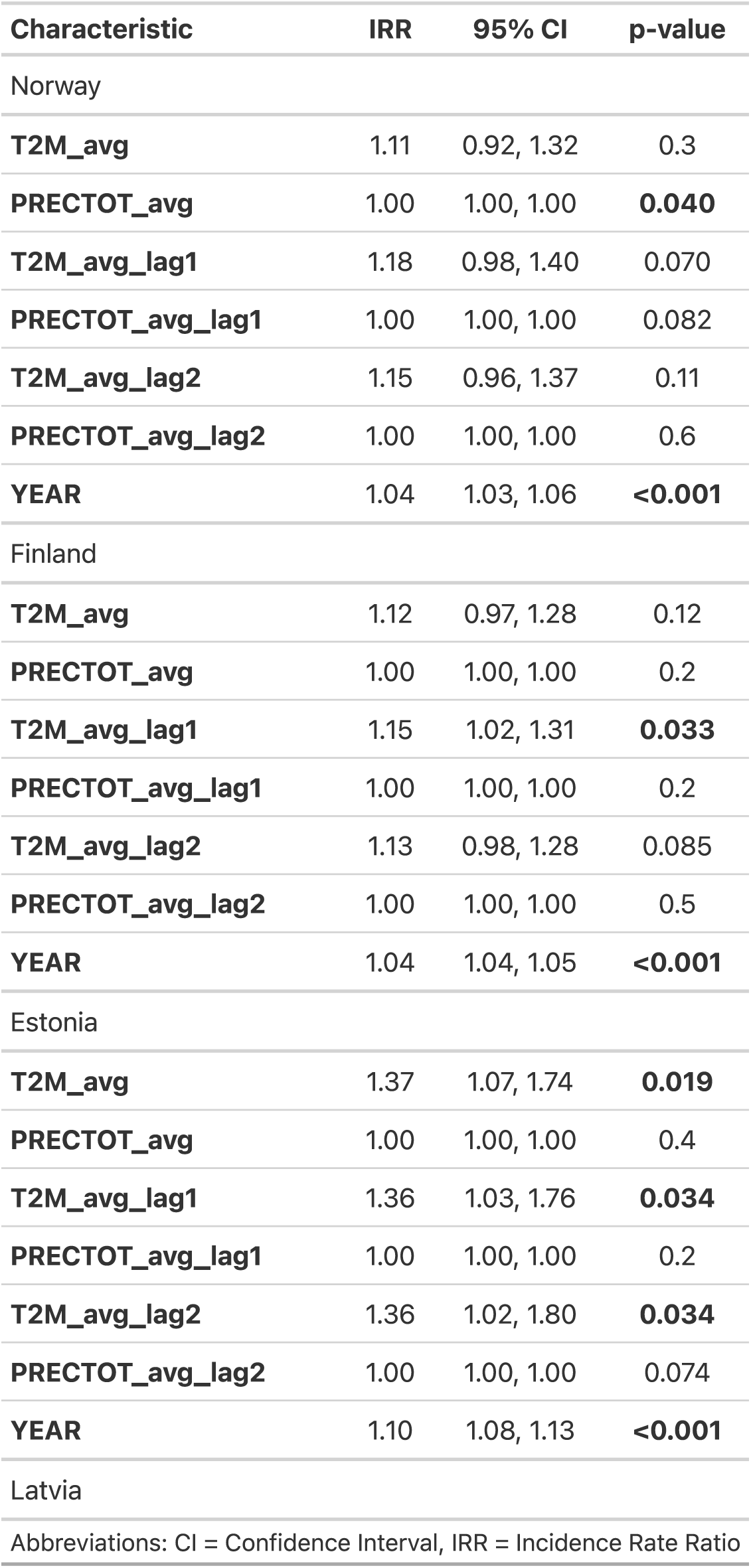
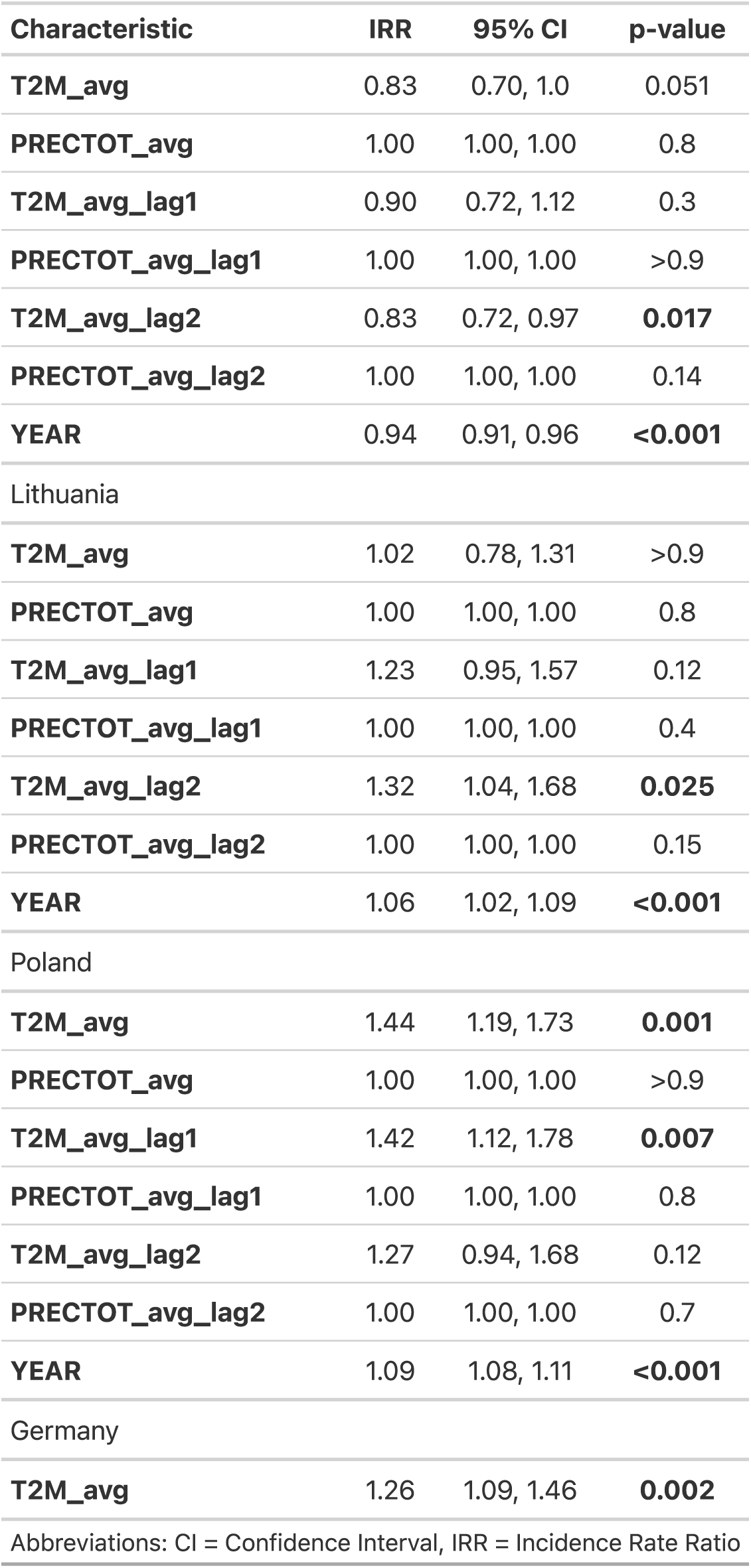
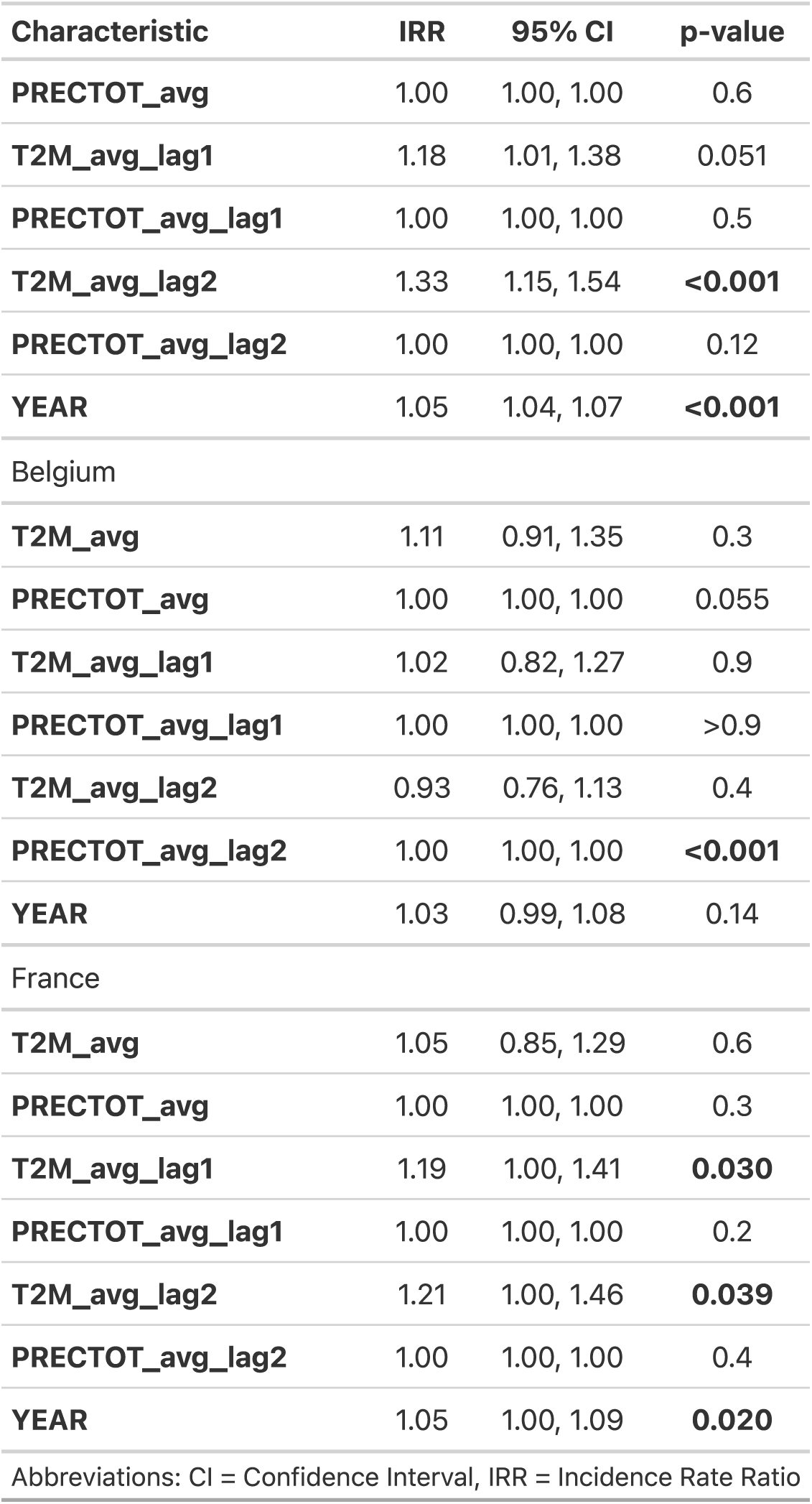
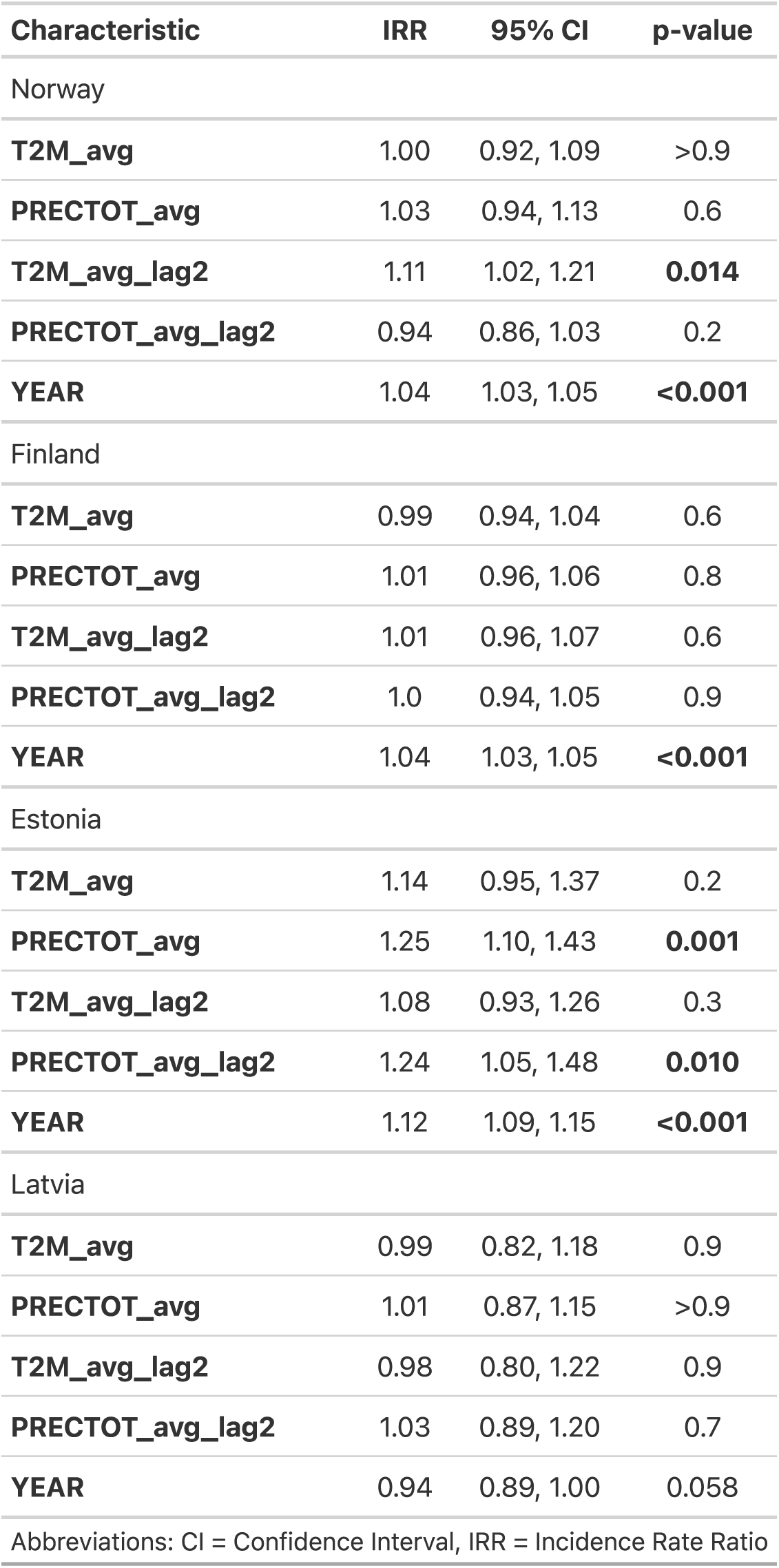
Complete Univariable Analysis.

| Characteristic | IRR | 95% CI | p-value |
| --- | --- | --- | --- |
| Norway |  |  |  |
| T2M_avg | 1.11 | 0.92, 1.32 | 0.3 |
| PRECTOT_avg | 1.00 | 1.00, 1.00 | **0.040** |
| T2M_avg_lag1 | 1.18 | 0.98, 1.40 | 0.070 |
| PRECTOT_avg_lag1 | 1.00 | 1.00, 1.00 | 0.082 |
| T2M_avg_lag2 | 1.15 | 0.96, 1.37 | 0.11 |
| PRECTOT_avg_lag2 | 1.00 | 1.00, 1.00 | 0.6 |
| YEAR | 1.04 | 1.03, 1.06 | **<0.001** |
| Finland |  |  |  |
| T2M_avg | 1.12 | 0.97, 1.28 | 0.12 |
| PRECTOT_avg | 1.00 | 1.00, 1.00 | 0.2 |
| T2M_avg_lag1 | 1.15 | 1.02, 1.31 | **0.033** |
| PRECTOT_avg_lag1 | 1.00 | 1.00, 1.00 | 0.2 |
| T2M_avg_lag2 | 1.13 | 0.98, 1.28 | 0.085 |
| PRECTOT_avg_lag2 | 1.00 | 1.00, 1.00 | 0.5 |
| YEAR | 1.04 | 1.04, 1.05 | **<0.001** |
| Estonia |  |  |  |
| T2M_avg | 1.37 | 1.07, 1.74 | **0.019** |
| PRECTOT_avg | 1.00 | 1.00, 1.00 | 0.4 |
| T2M_avg_lag1 | 1.36 | 1.03, 1.76 | **0.034** |
| PRECTOT_avg_lag1 | 1.00 | 1.00, 1.00 | 0.2 |
| T2M_avg_lag2 | 1.36 | 1.02, 1.80 | **0.034** |
| PRECTOT_avg_lag2 | 1.00 | 1.00, 1.00 | 0.074 |
| YEAR | 1.10 | 1.08, 1.13 | **<0.001** |
| Latvia |  |  |  |
| Abbreviations: CI = Confidence Interval, IRR = Incidence Rate Ratio |  |  |  |
| Norway |  |  |  |
| T2M_avg | 1.00 | 0.92, 1.09 | >0.9 |
| PRECTOT_avg | 1.03 | 0.94, 1.13 | 0.6 |
| T2M_avg_lag2 | 1.11 | 1.02, 1.21 | **0.014** |
| PRECTOT_avg_lag2 | 0.94 | 0.86, 1.03 | 0.2 |
| YEAR | 1.04 | 1.03, 1.05 | **<0.001** |
| Finland |  |  |  |
| T2M_avg | 0.99 | 0.94, 1.04 | 0.6 |
| PRECTOT_avg | 1.01 | 0.96, 1.06 | 0.8 |
| T2M_avg_lag2 | 1.01 | 0.96, 1.07 | 0.6 |
| PRECTOT_avg_lag2 | 1.0 | 0.94, 1.05 | 0.9 |
| YEAR | 1.04 | 1.03, 1.05 | **<0.001** |
| Estonia |  |  |  |
| T2M_avg | 1.14 | 0.95, 1.37 | 0.2 |
| PRECTOT_avg | 1.25 | 1.10, 1.43 | **0.001** |
| T2M_avg_lag2 | 1.08 | 0.93, 1.26 | 0.3 |
| PRECTOT_avg_lag2 | 1.24 | 1.05, 1.48 | **0.010** |
| YEAR | 1.12 | 1.09, 1.15 | **<0.001** |
| Latvia |  |  |  |
| T2M_avg | 0.99 | 0.82, 1.18 | 0.9 |
| PRECTOT_avg | 1.01 | 0.87, 1.15 | >0.9 |
| T2M_avg_lag2 | 0.98 | 0.80, 1.22 | 0.9 |
| PRECTOT_avg_lag2 | 1.03 | 0.89, 1.20 | 0.7 |
| YEAR | 0.94 | 0.89, 1.00 | 0.058 |
| Abbreviations: CI = Confidence Interval, IRR = Incidence Rate Ratio |  |  |  |

**Table A1: Complete Univariable Analysis**
| Characteristic | IRR | 95% CI | p-value |
| --- | --- | --- | --- |
| T2M_avg | 0.83 | 0.70, 1.0 | 0.051 |
| PRECTOT_avg | 1.00 | 1.00, 1.00 | 0.8 |
| T2M_avg_lag1 | 0.90 | 0.72, 1.12 | 0.3 |
| PRECTOT_avg_lag1 | 1.00 | 1.00, 1.00 | >0.9 |
| T2M_avg_lag2 | 0.83 | 0.72, 0.97 | **0.017** |
| PRECTOT_avg_lag2 | 1.00 | 1.00, 1.00 | 0.14 |
| YEAR | 0.94 | 0.91, 0.96 | **<0.001** |
| Lithuania |  |  |  |
| T2M_avg | 1.02 | 0.78, 1.31 | >0.9 |
| PRECTOT_avg | 1.00 | 1.00, 1.00 | 0.8 |
| T2M_avg_lag1 | 1.23 | 0.95, 1.57 | 0.12 |
| PRECTOT_avg_lag1 | 1.00 | 1.00, 1.00 | 0.4 |
| T2M_avg_lag2 | 1.32 | 1.04, 1.68 | **0.025** |
| PRECTOT_avg_lag2 | 1.00 | 1.00, 1.00 | 0.15 |
| YEAR | 1.06 | 1.02, 1.09 | **<0.001** |
| Poland |  |  |  |
| T2M_avg | 1.44 | 1.19, 1.73 | **0.001** |
| PRECTOT_avg | 1.00 | 1.00, 1.00 | >0.9 |
| T2M_avg_lag1 | 1.42 | 1.12, 1.78 | **0.007** |
| PRECTOT_avg_lag1 | 1.00 | 1.00, 1.00 | 0.8 |
| T2M_avg_lag2 | 1.27 | 0.94, 1.68 | 0.12 |
| PRECTOT_avg_lag2 | 1.00 | 1.00, 1.00 | 0.7 |
| YEAR | 1.09 | 1.08, 1.11 | **<0.001** |
| Germany |  |  |  |
| T2M_avg | 1.26 | 1.09, 1.46 | **0.002** |
| Abbreviations: CI = Confidence Interval, IRR = Incidence Rate Ratio |  |  |  |

**Table A1: Complete Univariable Analysis**
| Characteristic | IRR | 95% CI | p-value |
| --- | --- | --- | --- |
| PRECTOT_avg | 1.00 | 1.00, 1.00 | 0.6 |
| T2M_avg_lag1 | 1.18 | 1.01, 1.38 | 0.051 |
| PRECTOT_avg_lag1 | 1.00 | 1.00, 1.00 | 0.5 |
| T2M_avg_lag2 | 1.33 | 1.15, 1.54 | **<0.001** |
| PRECTOT_avg_lag2 | 1.00 | 1.00, 1.00 | 0.12 |
| YEAR | 1.05 | 1.04, 1.07 | **<0.001** |
| Belgium |  |  |  |
| T2M_avg | 1.11 | 0.91, 1.35 | 0.3 |
| PRECTOT_avg | 1.00 | 1.00, 1.00 | 0.055 |
| T2M_avg_lag1 | 1.02 | 0.82, 1.27 | 0.9 |
| PRECTOT_avg_lag1 | 1.00 | 1.00, 1.00 | >0.9 |
| T2M_avg_lag2 | 0.93 | 0.76, 1.13 | 0.4 |
| PRECTOT_avg_lag2 | 1.00 | 1.00, 1.00 | **<0.001** |
| YEAR | 1.03 | 0.99, 1.08 | 0.14 |
| France |  |  |  |
| T2M_avg | 1.05 | 0.85, 1.29 | 0.6 |
| PRECTOT_avg | 1.00 | 1.00, 1.00 | 0.3 |
| T2M_avg_lag1 | 1.19 | 1.00, 1.41 | **0.030** |
| PRECTOT_avg_lag1 | 1.00 | 1.00, 1.00 | 0.2 |
| T2M_avg_lag2 | 1.21 | 1.00, 1.46 | **0.039** |
| PRECTOT_avg_lag2 | 1.00 | 1.00, 1.00 | 0.4 |
| YEAR | 1.05 | 1.00, 1.09 | **0.020** |
| Abbreviations: CI = Confidence Interval, IRR = Incidence Rate Ratio |  |  |  |

**Table A2:**
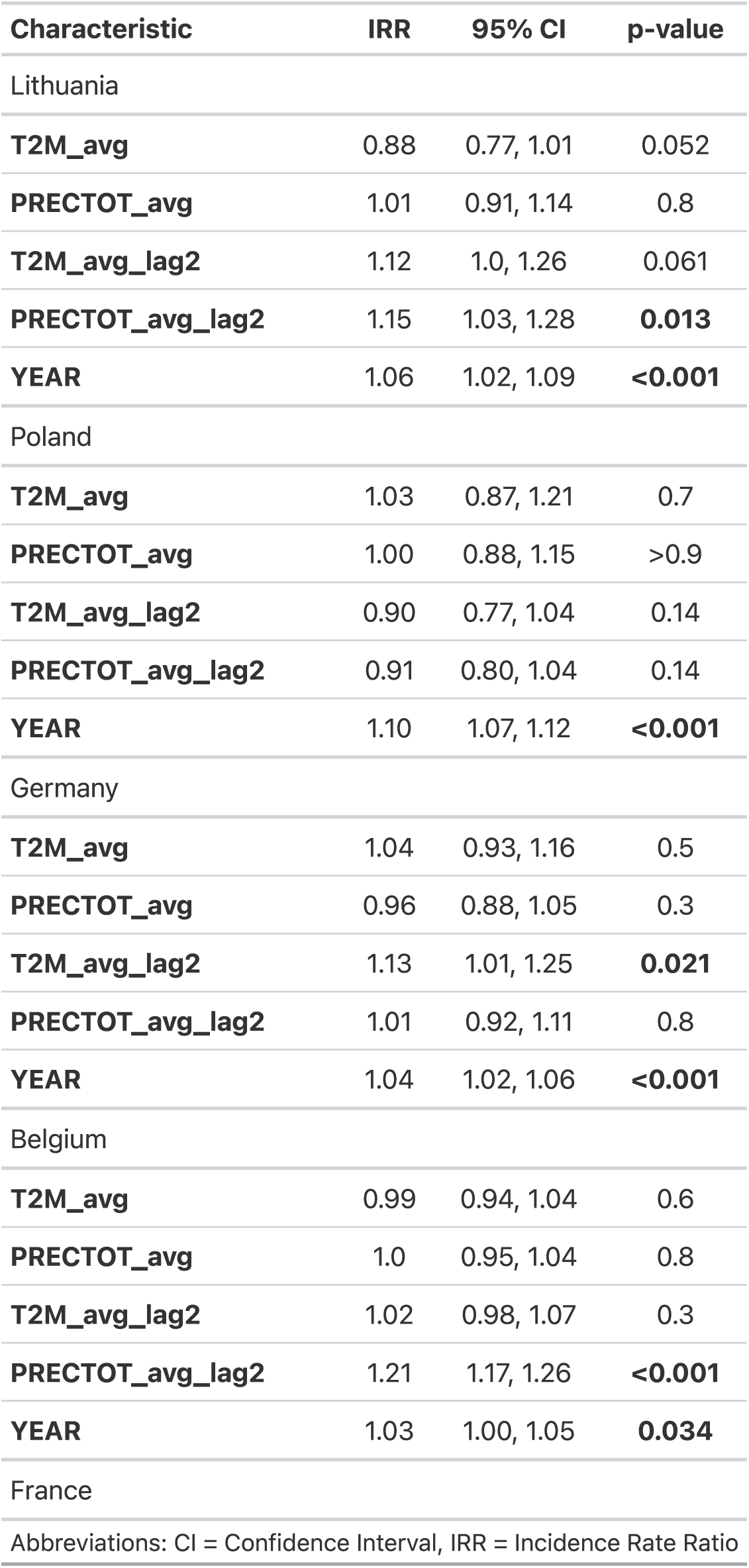
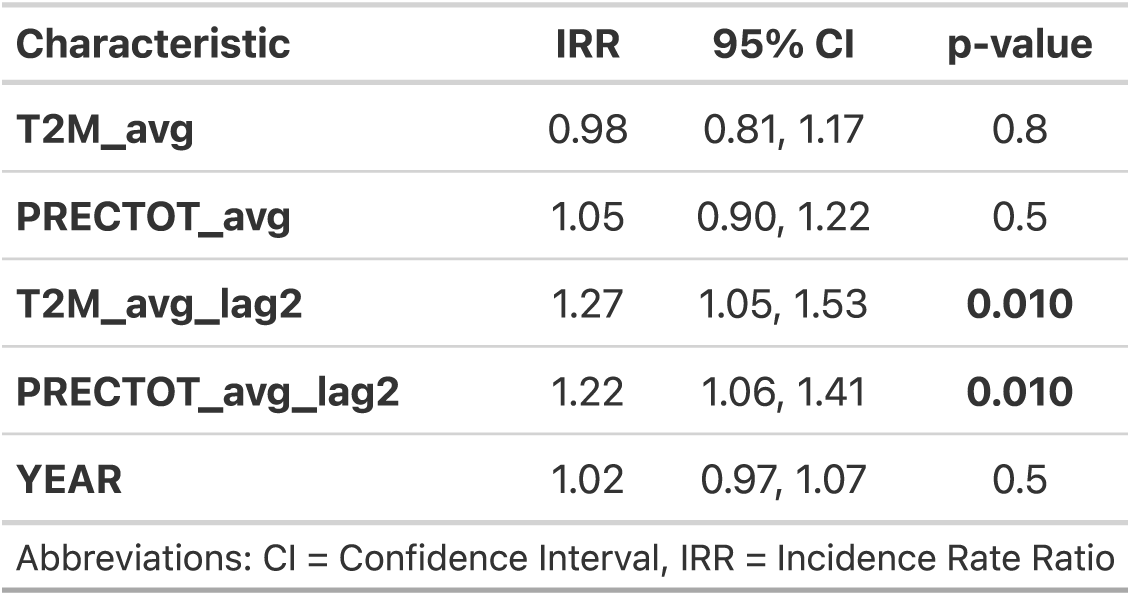
Complete Multivariable Analysis.

## Notes

### Competing Interest Statement

The authors have declared no competing interest.

### Funding Statement

This study did not receive any funding.

### Author Declarations

The study used ONLY openly available human data from: - Lyme disease incidence: Johns Hopkins Lyme Tracker (https://www.hopkinslymetracker.org)

### Summary of Updates

Revised for journal formatting and submission requirements.

